# Determinants of Radiation Exposure and Radiation Overexposure During Percutaneous Coronary Intervention A Real-World Study of 10,634 Procedures With Development of a Pragmatic Radiation-Alert Risk Score

**DOI:** 10.64898/2026.09.18.26363451

**Authors:** Franck Digne, Arthur Darmon, Victor Stratiev, Mohamed Abdellaoui, Julien Dreyfus, Maurizio Taramasso, Francesco Nappi, Mohammed Nejjari

**Affiliations:** Cardiology Department, Centre Cardiologique du Nord, Saint-Denis, France; Herzzentrum Hirslanden Zürich, Zürich, Switzerland; University of Zürich, Zürich, Switzerland

**Keywords:** percutaneous coronary intervention, radiation exposure, air kerma, chronic total occlusion, radioprotection, risk score

## Abstract

**Background:** Ionizing radiation is inherent to percutaneous coronary intervention (PCI) and may reach radiation-alert thresholds during complex procedures. Real-world data translating dose determinants into a usable planning-stage alert tool remain scarce.

**Aims:** To determine the prevalence and predictors of radiation overexposure (RO) during PCI and to develop a pragmatic radiation-alert risk score usable before substantial dose accumulates.

**Methods:** We analysed 10,634 consecutive PCI procedures performed between January 2008 and May 2018. Complex PCI was defined as unprotected left main, rotational atherectomy, chronic total occlusion (CTO), or bifurcation/trifurcation PCI. The composite RO endpoint was a dose-area product >500 Gy·cm², cumulative reference-point air kerma (AK) >5 Gy, or fluoroscopy time >60 minutes. A pragmatic score was built from early procedural variables.

**Results:** Overall, 3,457 procedures (32.5%) were complex, and RO occurred in 99 (0.93%). Complex PCI showed higher RO rates than standard PCI (2.1% vs 0.4%; p<0.001). In multivariable analysis, body mass index (BMI), male sex, CTO, rotational atherectomy, bifurcation, and treatment of ≥2 vessels independently predicted higher AK. A pragmatic score derived from early clinical and procedural variables showed good discrimination (area under the curve 0.772; 95% bootstrap CI 0.725–0.818) and was well calibrated (Brier score 0.0091; observed-to-expected ratio 0.994) for the composite RO endpoint.

**Conclusions:** Radiation-alert thresholds were exceeded in approximately 1% of PCI procedures, but fivefold more often during complex PCI. A pragmatic, internally assessed score based on early procedural variables identified high-risk cases with good discrimination, supporting planning-stage risk stratification to activate dose-sparing strategies before substantial radiation has accumulated.

**Clinical Perspective:** *What Is New?:* In 10,634 consecutive percutaneous coronary interventions, professional-society radiation-alert thresholds (dose-area product >500 Gy·cm², air kerma >5 Gy, or fluoroscopy time >60 minutes) were exceeded in 0.93% of procedures, five times more often during complex intervention, and the three metrics identified largely non-overlapping events. The RADAR-PCI score, an integer score based on seven characteristics available after diagnostic angiography, identified procedures at risk of radiation overexposure with good discrimination and calibration, and also stratified exceedance of the 2 Gy and 3 Gy air-kerma levels used for patient information.

*What Are the Clinical Implications?:* Because the score uses information available early in the procedure, radioprotective measures and structured post-procedural skin surveillance can be planned in advance for the minority of patients who need them most, rather than reactively once substantial dose has accumulated. A freely accessible online calculator (https://radar-pci.netlify.app) returns the score, the calibrated probability of exceeding each alert threshold, and model-based estimates of the expected dose metrics; external validation is warranted before broad adoption.

## Introduction

Percutaneous coronary intervention (PCI) is central to the management of acute coronary syndromes and symptomatic obstructive coronary artery disease. Contemporary guidelines and advances in devices, intracoronary imaging, and operator expertise have expanded PCI to increasingly complex anatomical subsets.^1,2^ While these developments have improved clinical outcomes, they have also increased radiation exposure because complex PCI often requires prolonged fluoroscopy, multiple angiographic acquisitions, adjunctive imaging, and longer procedural duration.^3^

Radiation exposure during PCI raises concerns regarding both stochastic effects, including malignancy, and deterministic effects such as erythema, epilation, tissue injury, and chronic radiodermatitis.^4–6^ In clinical practice, radiation burden is commonly assessed using dose-area product (DAP), cumulative reference-point air kerma (AK), and fluoroscopy time. DAP reflects overall radiation energy delivered, whereas AK is more closely related to potential skin injury; fluoroscopy time remains a less reliable surrogate when other dose metrics are available.^5,7^ Professional societies have established operational alert thresholds and post-procedural surveillance strategies for high-dose fluoroscopically guided interventions.^5–7^

Such recommendations are particularly relevant because radiation-induced skin injury may occur weeks or months after exposure and remains frequently underrecognized.^8,9^ Large contemporary registries have shown that radiation exposure during PCI has decreased over time but remains highly variable across institutions and procedures.^10^ Determinants of radiation burden include patient characteristics, lesion complexity, and operator practice patterns.^11^ Cutaneous radiation injury after repeated coronary interventions further underscores the relevance of radiation monitoring.^12,13^ High radiation-dose procedures remain common, supporting operational dose-alert thresholds.^14^ Patient body habitus, particularly BMI, is consistently associated with increased patient and operator dose.^15–17^ The institutional frequency of high-dose procedures, including those exceeding 5 Gy AK, varies substantially.^18^

Despite these advances, important gaps remain. Most studies describe radiation distributions rather than actionable alert thresholds, few integrate DAP, AK, and fluoroscopy time into a composite endpoint, and bedside prediction of radiation overexposure remains poorly developed. We therefore analysed a large real-world PCI cohort to determine the prevalence and predictors of radiation overexposure and to develop a pragmatic radiation-alert score applicable after diagnostic angiography and before intervention.

## Methods

### Study design and population

This retrospective observational cohort study used the anonymized institutional database of the Department of Interventional Cardiology, Centre Cardiologique du Nord, including all consecutive patients who underwent PCI between January 2, 2008 and May 7, 2018. After database cleaning, harmonization of variable labels, and quality control of radiation metrics, the final analytical cohort included 10,634 PCI procedures. The study analyzed procedural-level radiation burden rather than patient-level cumulative exposure across repeated interventions; all analyses were performed on deidentified data. Throughout the study period, the catheterization laboratory comprised two dedicated rooms with the same fluoroscopic systems, which underwent regular quality-control and dosimetric calibration but were not replaced, and all procedures were performed by four senior interventional cardiologists. This stability reduces the likelihood that variability in recorded radiation metrics reflected changes in hardware or operator experience. Fluoroscopy and cine acquisition were performed at 15 frames per second throughout the study period; acquisition settings were common to the two rooms and to all procedures, and no dedicated low-dose protocol for CTO or other complex PCI was in use.

### Definition of standard and complex PCI

Procedures were classified as complex PCI if at least one of the following predefined criteria was present: unprotected left main PCI, PCI requiring rotational atherectomy, CTO PCI, or bifurcation/trifurcation PCI. Procedures without any of these criteria were classified as standard PCI. CTO was defined as a coronary occlusion of presumed duration greater than 3 months; severe calcification as angiographic calcification of the treated proximal segment graded moderate-to-important or massive; and bifurcation/trifurcation PCI as treatment of a lesion involving a bifurcation or trifurcation. This definition was chosen because these lesion/procedure types are routinely recognized as technically demanding and have previously been associated with higher fluoroscopy time, contrast use, and radiation exposure.^10,11,14^

### Radiation metrics

The available radiation metrics were DAP (Gy·cm²), cumulative reference-point AK (mGy), and fluoroscopy time (seconds). DAP was available for almost all procedures. AK was available in 5,001 procedures after the metric became recorded in the institutional database and was analyzed both as a continuous endpoint in the AK-available cohort and as one component of the composite endpoint. Fluoroscopy time was available in 3,077 procedures. Dose metrics were recorded per procedural session: for ad hoc PCI they therefore include the diagnostic acquisition performed in the same session, whereas for staged PCI they cover the interventional session only. PCI was ad hoc in 9,205 procedures (86.6%) and staged in 1,429 (13.4%), staged procedures being identified through a recorded indication of prior diagnostic coronary angiography; 660 procedures (6.2%) were classified as urgent, a finer elective/urgent/emergent gradation not being recorded. Procedure duration and contrast volume were analyzed as procedural descriptors. To assess the representativeness of the AK subcohort, baseline characteristics were compared between procedures with and without available AK: the two groups were well matched for age, BMI, comorbidities, and all measures of lesion complexity (all p>0.05). Because automated AK recording was implemented progressively, procedures with AK were performed between 2014 and 2018 and showed higher radial access, lower median DAP, and shorter duration, consistent with contemporaneous improvements in technique rather than differences in case mix.

### Radiation overexposure endpoint

RO was defined as the occurrence of at least one available Society of Interventional Radiology (SIR) radiation-alert threshold: DAP >500 Gy·cm², cumulative AK >5 Gy, or fluoroscopy time >60 minutes, assessed among the metrics recorded for each procedure, ^(5)^. Peak skin dose >3 Gy is also included in SIR thresholds but was not available in the institutional database and therefore could not be incorporated. RO denotes operational alert-threshold exceedance, not demonstrated tissue injury. The composite endpoint was selected because each available threshold is actionable and can trigger documentation, patient information, and surveillance.

### Data cleaning and quality control

Before analysis, the database underwent systematic quality control of radiation metrics and anthropometric variables. Five DAP or AK values were identified as isolated factor of 1,000 data-entry errors on source-record review and were corrected to their verified values; none represented a true alert-threshold exceedance. Body mass index was recomputed from height and weight after detection of a height/weight column inversion affecting 144 records, which were corrected by reinstating the correct orientation; an additional 24 physiologically impossible values that could not be resolved against the source record were set to missing. No patient or procedure was excluded on the basis of these corrections, and no value was imputed or inferred. Radiation metrics that were not recorded during the corresponding period were treated as missing and handled by available-case analysis, as detailed below.

### Statistical analysis

Continuous variables are presented as median [interquartile range] and compared using the Mann-Whitney U test; categorical variables as counts (percentages) and compared using the χ² or Fisher exact test. Because cumulative AK was right-skewed, it was log-transformed and analyzed by multivariable linear regression, with results expressed as the adjusted percentage change in AK with 95% confidence intervals. Procedures with available AK were also analyzed by AK quartiles. A pragmatic integer risk score for the composite RO endpoint was constructed from variables available early during the procedure. Candidate variables were selected on the basis of established determinants of procedural radiation reported in the literature and confirmed in our multivariable analysis, and point weights were assigned to reflect the relative strength of these associations and simplified to small integers to ensure bedside usability. Because the score was derived within the study cohort, it should be regarded as exploratory pending external validation. Score discrimination was assessed by the area under the receiver-operating characteristic curve (AUC), with a 95% confidence interval from a 1,000-replication bootstrap. A LASSO-penalized logistic regression with 10-fold cross-validation, using the same candidate variables, served as a benchmark. Model calibration was evaluated using the Brier score, calibration slope, and observed-to-expected ratio. Variance inflation factors assessed multicollinearity. Temporal trends were assessed using the Spearman rank correlation. A prespecified secondary analysis evaluated score discrimination for the AK >5 Gy threshold alone. A two-sided p<0.05 defined statistical significance. Analyses were performed in Python 3.11 (pandas, NumPy, SciPy, scikit-learn). DAP-only and Firth penalised-logistic sensitivity analyses, with optimism-corrected discrimination and operating characteristics, are detailed in the Supplementary Methods. The corresponding author had full access to all the data in the study and takes responsibility for its integrity and the data analysis.

### Ethics

This study was conducted in accordance with the Declaration of Helsinki and was approved by the Institutional Review Board of the Adène Fonds de dotation (IRB accreditation 0990-0279; approval number IRB_ADENE_20240902, 5 September 2024). The study was registered on ClinicalTrials.gov (NCT06565793). It was a retrospective analysis of de-identified data collected during routine clinical care, with no intervention on patients. Because the analysis used only anonymized, pre-existing data and entailed no modification of patient management, the requirement for written informed consent was waived by the Institutional Review Board, in accordance with applicable data protection regulations.

## Results

### Population and radiation-metric availability

The final cohort included 10,634 PCI procedures. Overall, 3,457 procedures (32.5%) met at least one complex PCI criterion and 7,177 (67.5%) were classified as standard PCI. DAP was available in 10,630 procedures (99.96%), cumulative AK in 5,001 procedures (47.0%), and fluoroscopy time in 3,077 procedures (28.9%) (Supplementary Table 1, Figure 1).

**Figure 1.**
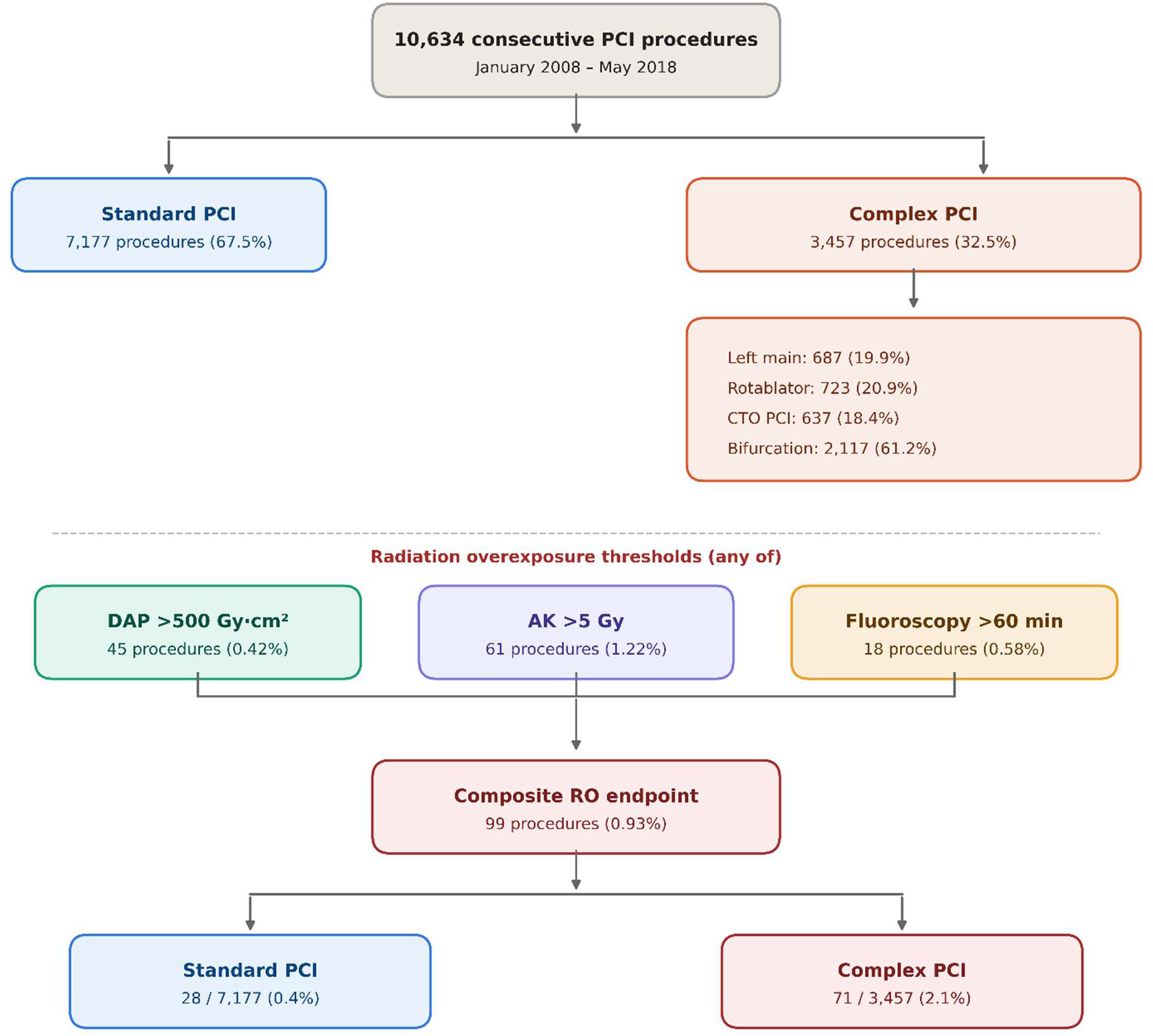
Study cohort, radiation-metric availability, and radiation overexposure endpoint. Flowchart showing derivation of the analytical cohort (10,634 consecutive PCI procedures). Availability of each radiation metric is depicted (DAP in 99.96%, AK in 47.0%, fluoroscopy time in 28.9%). The composite RO endpoint is decomposed into its three component thresholds (DAP >500 Gy·cm², AK >5 Gy, fluoroscopy time >60 min). AK = cumulative reference-point air kerma; DAP = dose-area product; PCI = percutaneous coronary intervention; RO = radiation overexposure.

### Prevalence of radiation overexposure

Overall, 99 procedures (0.93%) reached at least one available SIR radiation-alert threshold. Individual thresholds were exceeded in 45 procedures (0.42%) for DAP >500 Gy·cm², 61 procedures (0.57% of the total cohort; 1.22% of procedures with AK available) for AK >5 Gy, and 18 procedures (0.17% of the total cohort; 0.58% of procedures with fluoroscopy time available) for fluoroscopy time >60 minutes. The thresholds overlapped incompletely: 30 cases reached the DAP threshold only, 42 reached the AK threshold only, 8 reached the fluoroscopy-time threshold only, 9 met both DAP and AK thresholds, 4 met AK and fluoroscopy thresholds, and 6 reached all three available thresholds. This incomplete overlap confirms that DAP, AK, and fluoroscopy time are related but not interchangeable dose indicators (Supplementary Figure 1).

### Baseline characteristics

Patients undergoing complex PCI were slightly older. BMI and the prevalence of BMI ≥30 kg/m² were similar between groups. Complex PCI patients had lower creatinine clearance, more renal failure, more prior CABG, and more frequent femoral access. Dyslipidemia was also more frequent in the complex PCI group, whereas diabetes and current smoking were slightly less frequent (Supplementary Table 2).

### Procedural characteristics and radiation exposure

Complex PCI procedures were associated with a substantially higher procedural and radiation burden than standard PCI across all metrics examined, including procedure duration, fluoroscopy time, DAP, cumulative AK, and contrast volume (all p<0.001; Supplementary Table 3, Figure 2). Composite RO was more frequent after complex PCI, occurring in 71/3,457 (2.1%) versus 28/7,177 standard PCI procedures (0.4%; p<0.001), a fivefold difference.

**Figure 2.**
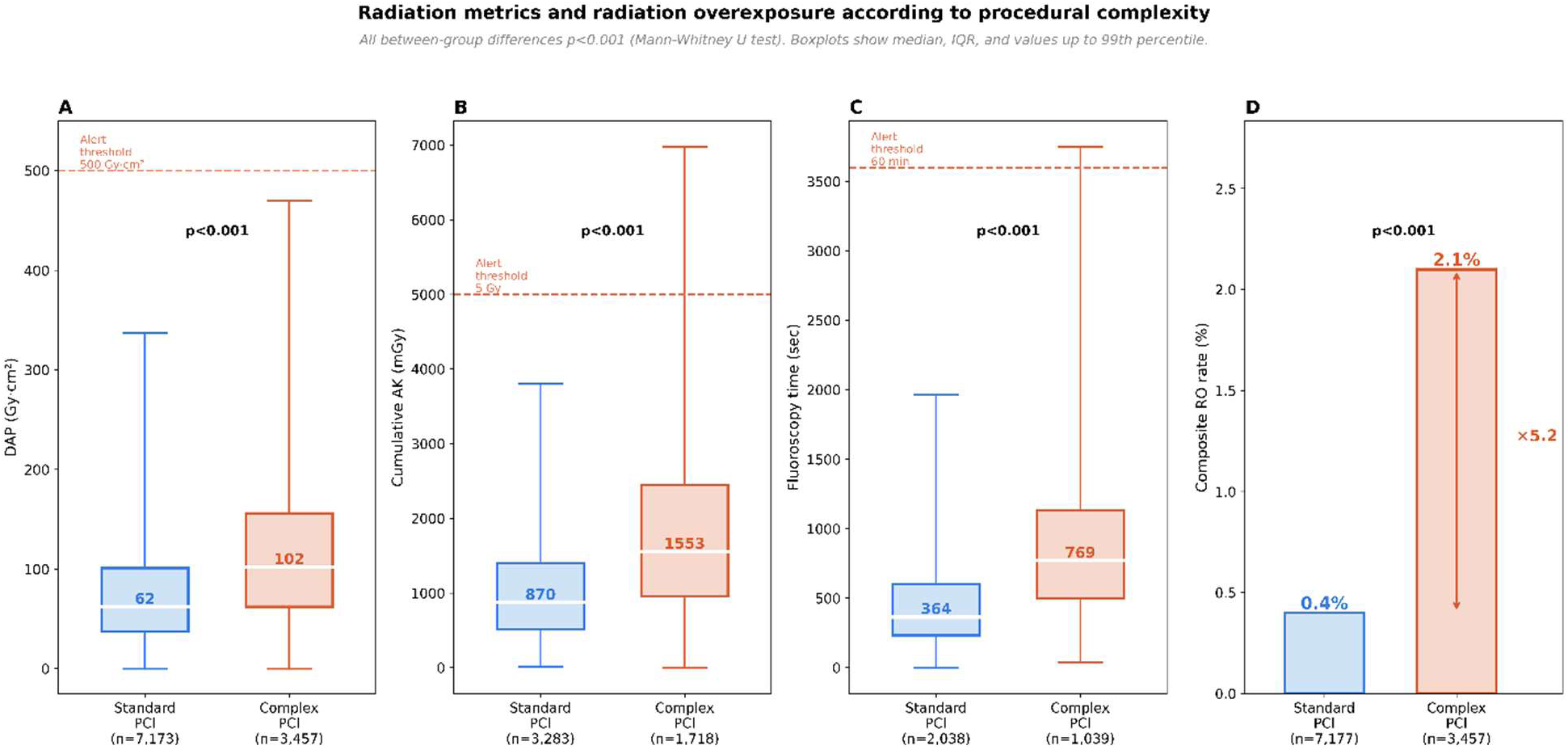
Radiation metrics and radiation overexposure according to procedural complexity. Box plots (median, interquartile range, and outliers) comparing DAP (Gy·cm²), AK (mGy), and fluoroscopy time (seconds) between standard PCI (n=7,177) and complex PCI (n=3,457). Inset bar chart shows composite RO rate (0.4% vs 2.1%; p<0.001). All between-group differences p<0.001 by Mann-Whitney U test. AK = cumulative reference-point air kerma; DAP = dose-area product; PCI = percutaneous coronary intervention; RO = radiation overexposure.

### Radiation burden across AK quartiles

In the AK-available cohort, higher AK quartiles showed a graded increase in radiation and procedural burden, with median AK, DAP, procedure duration, fluoroscopy time, and contrast volume all rising progressively from the first to the fourth quartile (all p<0.001; Supplementary Table 4). Obesity, complex PCI, CTO PCI, rotational atherectomy, bifurcation PCI, and ≥2 treated vessels likewise increased progressively across quartiles (Supplementary Figure 2). Composite RO was essentially restricted to the highest AK quartile, where it occurred in 70/1,250 procedures (5.6%).

### Predictors of increasing AK

In the multivariable log(AK) model, BMI was a strong independent patient-related determinant of higher AK, with each 5-kg/m² increase associated with a +35.1% relative increase (95% CI +32.2% to +38.1%; p<0.001), as was male sex (+35.6%; p<0.001). The strongest procedural predictors were rotational atherectomy (+89.6%) and CTO PCI (+87.9%), followed by bifurcation/trifurcation PCI (+45.0%), treatment of ≥2 vessels excluding the left main (+33.5%), and left main PCI (+18.2%) (all p<0.001). Femoral access was associated with a smaller but significant increase (+6.3%; p=0.044), whereas severe calcification was associated with lower AK (-18.7%; p<0.001). Age, diabetes, and renal failure were not independently associated with AK (Table 1).

**Table 1.** Multivariable predictors of increased cumulative reference-point air kerma (log-transformed AK model)

| Predictor | Relative change in AK, % | 95% CI | p-value |
| --- | --- | --- | --- |
| Age per 10 years | +0.5 | -1.3 to +2.3 | 0.586 |
| Male sex | +35.6 | +29.4 to +42.2 | <0.001 |
| BMI per 5 kg/m<sup>2</sup> | +35.1 | +32.2 to +38.1 | <0.001 |
| Femoral access | +6.3 | +0.2 to +12.8 | 0.044 |
| Diabetes | +3.0 | -1.1 to +7.3 | 0.152 |
| Renal failure | -6.0 | -14.1 to +2.9 | 0.183 |
| CTO PCI | +87.9 | +72.7 to +104.6 | <0.001 |
| Rotational atherectomy | +89.6 | +76.3 to +103.9 | <0.001 |
| Bifurcation/trifurcation PCI | +45.0 | +38.0 to +52.4 | <0.001 |
| Left main PCI | +18.2 | +7.7 to +29.7 | <0.001 |
| ≥2 vessels treated (left main counted as one vessel) | +33.5 | +23.0 to +45.0 | <0.001 |
| Severe calcification | -18.7 | -22.2 to -14.9 | <0.001 |

Results are adjusted relative percentage change in AK per unit change in each predictor, derived from multivariable linear regression on log-transformed AK. AK = cumulative reference-point air kerma; BMI = body mass index; CI = confidence interval; CTO = chronic total occlusion; PCI = percutaneous coronary intervention.

### Radiation overexposure risk score

The resulting score, designated RADAR-PCI (Radiation Alert Dose Assessment Risk score), assigned 1 point for BMI 30-34.9 kg/m², 2 points for BMI ≥35 kg/m², 1 point for femoral access, 2 points for treatment of ≥2 vessels (the left main, when treated, counting as one vessel), 1 point for severe calcification, 2 points for bifurcation/trifurcation PCI, 3 points for rotational atherectomy, and 4 points for CTO PCI (Table 2). Risk categories were defined as low risk (0-2 points), intermediate risk (3-5 points), and high risk (≥6 points). These category thresholds were chosen for clinical usability after inspection of the score distribution and were not optimized against the endpoint. RO increased progressively from 31/7,572 (0.41%) in the low-risk group to 40/2,397 (1.67%) in the intermediate-risk group and 28/665 (4.21%) in the high-risk group (Table 3, Supplementary Figure 3). The simplified score had an AUC of 0.772 (95% bootstrap CI 0.725–0.818; 1,000 replications) for predicting the composite endpoint. A penalized logistic model using the same candidate predictors, fitted with L1 regularisation (LASSO) and λ selected by tenfold cross-validation, had an in-sample AUC of 0.778 (cross-validated AUC 0.764), supporting the clinical informativeness of the selected factors while confirming that model-derived estimates may be optimistic in rare-event settings (Table 4, Figure 3). At a threshold of 3 points the score had 68.7% sensitivity and 99.6% negative predictive value, and at 6 points 94.0% specificity (Supplementary Table 5). The corresponding positive predictive values were 2.2% (95% CI 1.8 to 2.8) at ≥3 points and 4.2% (2.9 to 6.0) at ≥6 points, with positive likelihood ratios of 2.42 and 4.68, a profile of risk enrichment rather than rule-in prediction.

**Figure 3.**
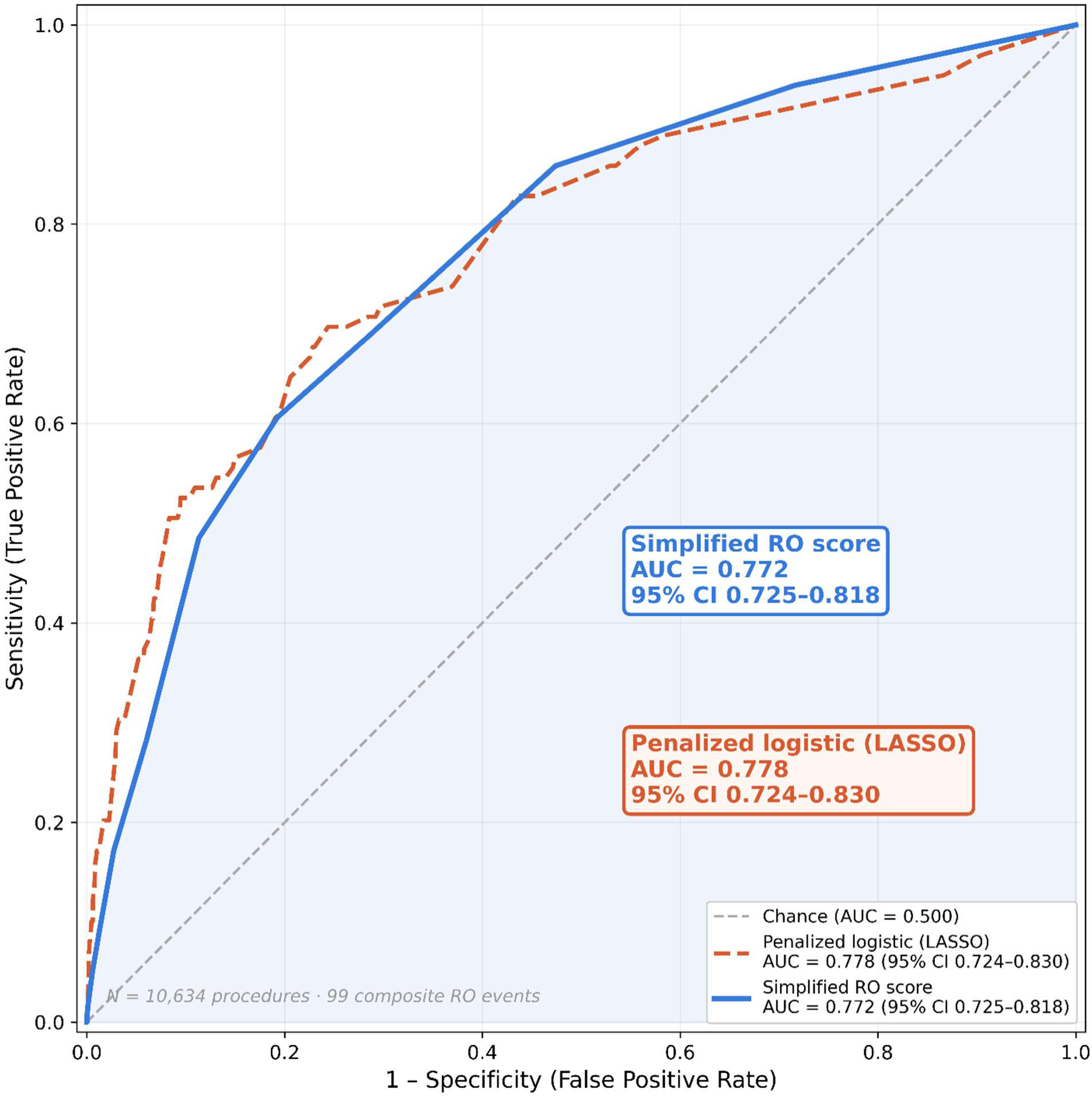
Receiver-operating characteristic curve for the simplified radiation overexposure risk score. ROC curves for the simplified RO risk score (AUC 0.772, solid line) and the penalized logistic model, LASSO with tenfold cross-validation (in-sample AUC 0.778, cross-validated AUC 0.764, dashed line) predicting the composite radiation-alert endpoint. The diagonal reference line represents chance discrimination. AK = cumulative reference-point air kerma; AUC = area under the receiver-operating characteristic curve; CV = cross-validated; DAP = dose-area product; LASSO = least absolute shrinkage and selection operator; RO = radiation overexposure; ROC = receiver-operating characteristic.

**Table 2.**
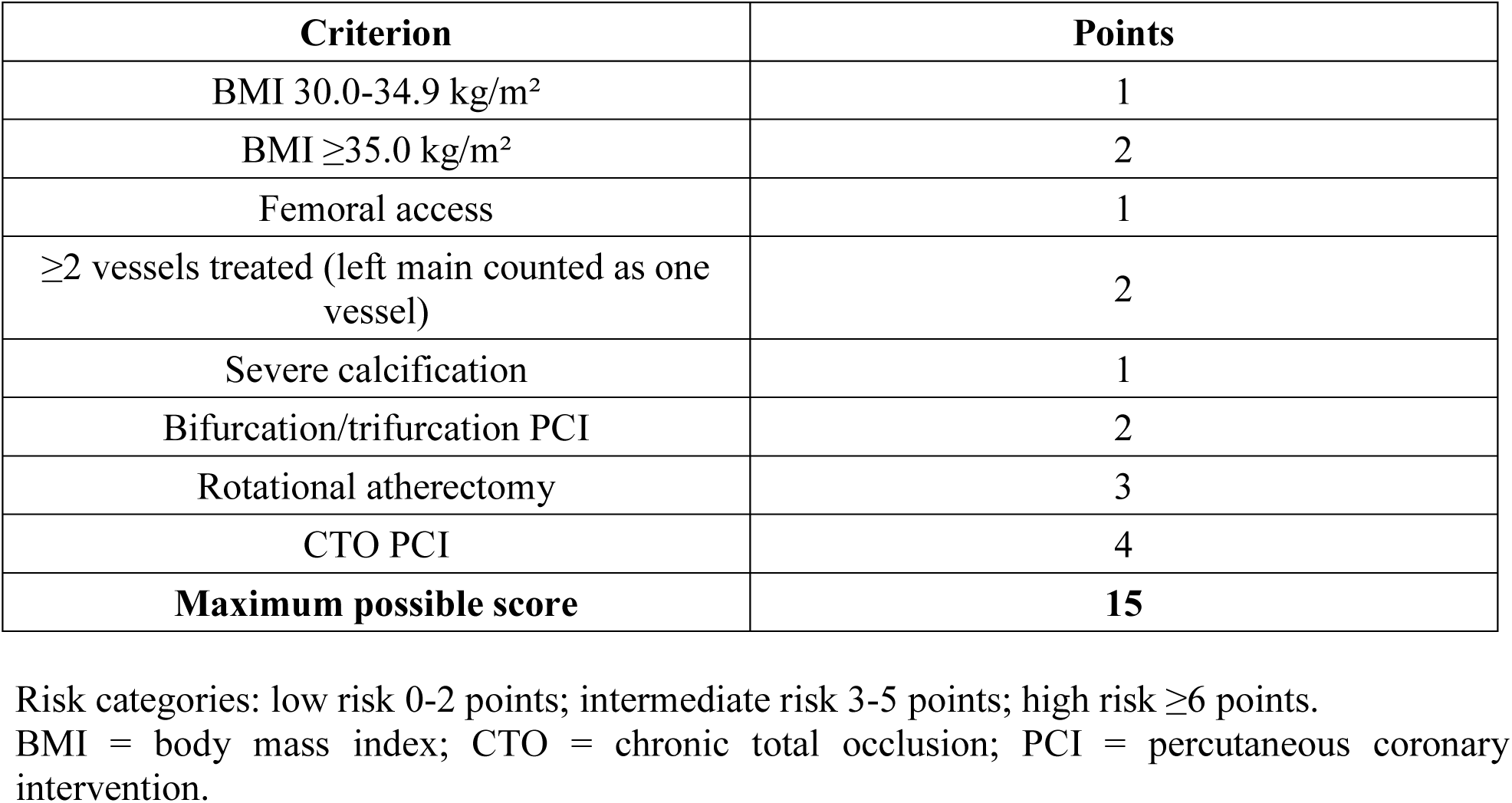
Components of the radiation overexposure risk score.

| **Criterion** | **Points** |
| --- | --- |
| BMI 30.0-34.9 kg/m<sup>2</sup> | 1 |
| BMI ≥35.0 kg/m<sup>2</sup> | 2 |
| Femoral access | 1 |
| ≥2 vessels treated (left main counted as one vessel) | 2 |
| Severe calcification | 1 |
| Bifurcation/trifurcation PCI | 2 |
| Rotational atherectomy | 3 |
| CTO PCI | 4 |
| **Maximum possible score** | **15** |

Risk categories: low risk 0-2 points; intermediate risk 3-5 points; high risk ≥ 6 points.
BMI = body mass index; CTO = chronic total occlusion; PCI = percutaneous coronary intervention.

**Table 3.**
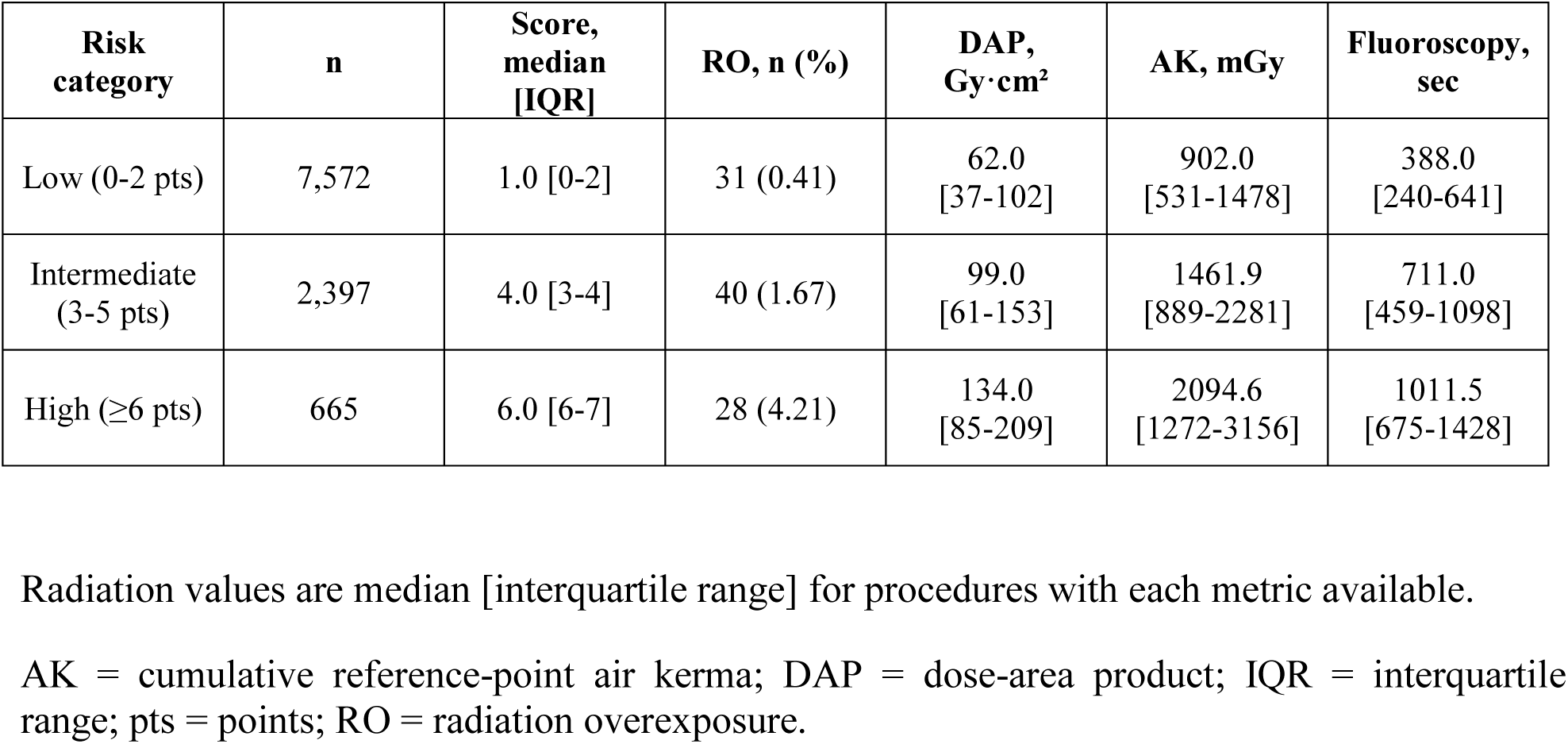
Observed radiation overexposure rate and radiation metrics by score risk category.

**Table 4.** Discrimination of the simplified radiation overexposure risk score and the penalised multivariable model.

| Model | AUC (95% CI) |
| --- | --- |
| Simplified RO risk score | 0.772 (0.725-0.818) |
| Penalised logistic (LASSO) | 0.778 (CV: 0.764) |
| Score, AK >5 Gy endpoint (secondary) | 0.816 (0.763-0.863) |

AUC = area under the receiver-operating characteristic curve; CV = cross-validated; RO = radiation overexposure. LASSO = least absolute shrinkage and selection operator.
Brier score 0.0091; observed/expected ratio 0.994. Secondary analysis on n=5,001 with AK available.

The score was well calibrated, with a Brier score of 0.0091, a calibration slope of 0.96, and an observed-to-expected event ratio of 0.994. Observed and predicted RO rates were concordant across categories (low risk, 0.41% observed vs 0.45% predicted; intermediate, 1.67% vs 1.55%; high, 4.21% vs 4.95%) (Supplementary Table 6). All variance inflation factors were below 1.6 (severe calcification, 1.19), indicating no meaningful multicollinearity. Complexity criteria overlapped only modestly: among the 3,457 complex procedures, 2,824 (81.7%) met a single criterion, 559 (16.2%) met two, and 74 (2.1%) met three, and the combination of CTO with rotational atherectomy was rare (n=24) (Supplementary Table 9). The negative adjusted coefficient for severe calcification reflected its conditional association with rotational atherectomy (86% of rotational atherectomy procedures were severely calcified) rather than collinearity, and a sensitivity analysis excluding rotational atherectomy and CTO PCI attenuated this coefficient toward the null (Supplementary Table 7). A further sensitivity analysis removing severe calcification from the score gave similar discrimination (AUC 0.780, 95% bootstrap CI 0.730 to 0.826; Supplementary Table 8); the criterion was retained because calcification is directly observable on the diagnostic angiogram, whereas rotational atherectomy is sometimes decided only during the procedure, so the calcification point preserves risk information when atherectomy has not yet been decided.

Across the study period, median DAP declined from 127 Gy·cm² in 2008 to 64 Gy·cm² in 2018 (Spearman rho = - 0.22, p<0.001), consistent with the broader temporal trend toward lower radiation exposure reported in contemporary PCI cohorts and with increasing radiation-protection awareness, while equipment and operator composition remained stable. Cumulative AK showed no temporal trend, as automated AK recording was introduced only in the later study years (0% of procedures before 2014, 100% from 2016 onward), reflecting a technological change rather than random missingness.

In a prespecified secondary analysis restricted to the 5,001 procedures with available AK, the score discriminated the AK >5 Gy threshold alone with an AUC of 0.816 (95% CI 0.763–0.863), exceeding its discrimination for the composite endpoint. Discrimination was preserved for the DAP-only endpoint ascertained cohort-wide (AUC 0.716, 95% CI 0.637–0.798), and the optimism-corrected AUC for the composite endpoint was 0.773. Within the AK subcohort the composite endpoint occurred in 71 procedures (1.42%) with an AUC of 0.801 (95% bootstrap CI 0.751 to 0.849), and among the 3,077 procedures with all three metrics available, in 47 (1.53%) with an AUC of 0.803 (0.741 to 0.860), indicating that discrimination was preserved under uniform ascertainment. The score also stratified the substantial-dose levels used by many centres for patient information: air kerma above 2 Gy occurred in 968 procedures (19.4%) with an AUC of 0.724 (0.705 to 0.741), and above 3 Gy in 365 (7.3%) with an AUC of 0.752 (0.726 to 0.779) (Supplementary Table 8).

To support bedside use, the RADAR-PCI score was implemented as a freely accessible, self-contained online calculator (https://radar-pci.netlify.app). For any combination of the seven planning-stage criteria, the calculator returns the integer score (0 to 15), the corresponding risk category with its observed overexposure rate, and the calibrated probability of exceeding each of the three alert thresholds, together with model-based estimates of the expected dose-area product, cumulative air kerma and fluoroscopy time with 95% confidence intervals. All coefficients are embedded in the application and were verified programmatically against the analytical dataset; no patient data are transmitted or stored. The calculator is intended as a planning aid and does not replace real-time dose monitoring or clinical judgement.

## Discussion

In this large real-world cohort, radiation-alert threshold exceedance was uncommon (∼1%) but frequent enough to justify a structured management strategy. It was driven by the interaction of patient habitus and procedural complexity rather than any single factor, and a simple bedside score achieved good discrimination while retaining clinical interpretability.

A key contribution of this analysis is the use of a clinically grounded composite endpoint rather than a percentile-based definition that does not correspond to patient-management thresholds. The endpoint is built from established SIR, NCRP, and ICRP alert levels, all emphasizing predefined thresholds to trigger patient information, documentation, and surveillance.^(5,19–23)^

Similar analyses of procedures exceeding 5 Gy AK have shown substantial inter-institutional variability.^(14,18)^

Threshold exceedance does not prove skin injury, but because deterministic injury may appear weeks to months later, risk communication and post-procedural surveillance remain important.(20,22,23)

The incomplete overlap between DAP, AK, and fluoroscopy time is also clinically important: DAP reflects total energy over the irradiated field and is useful for stochastic risk, AK more closely approximates reference-point energy and is more relevant to deterministic skin effects, and fluoroscopy time ignores several acquisition factors. Because these indicators cannot be interconverted for individual management, dose surveillance should record all available metrics rather than a single surrogate.

Our findings align with the PROTECTION VIII study, the largest radiation survey in PCI to date. That study demonstrated a substantial decline in PCI-related radiation exposure over a decade but also persistent intersite variability and independent radiation predictors including coronary occlusion, left main PCI, and multivessel PCI.(10) The present cohort extends those observations by including detailed lesion subsets such as CTO, bifurcation PCI, and rotational atherectomy, while also anchoring the endpoint to SIR alert thresholds. The 88% relative AK increase with CTO PCI is biologically plausible, reflecting dual injection, guidewire escalation, dissection/re-entry techniques, and prolonged fluoroscopy.(24–26) A recent multicenter CTO registry reported a 52% reduction in air kerma over the past decade, with excess radiation (AK >5 Gy) falling from 15.8% to 3.7% of procedures, driven by equipment updates and risk awareness, although interoperator variability persisted.^(25)^ Our cohort reflects the same secular decline in procedural dose, supporting the contemporary relevance of these findings; however, whereas that improvement was driven largely by equipment upgrades, the residual interoperator variability they describe is precisely what a planning-stage alert score targets, by prompting dose-sparing behaviour before irradiation accumulates, independently of the imaging system.

Rotational atherectomy was another strong determinant of AK, reflecting its use in heavily calcified lesions requiring multiple runs and adjunctive imaging. The inverse adjusted association of severe calcification with AK should not be read as protective; it reflects the conditional structure of the model, the most radiation-intensive calcified disease being captured by atherectomy, CTO, and bifurcation rather than true collinearity. Severe calcification was nonetheless retained as a low-weight, readily available score component, pending prospective testing.

BMI emerged as one of the strongest patient-level determinants of radiation burden. Each 5- kg/m² increase was associated with a 35% increase in AK, consistent with prior work showing that patient size is a major driver of radiation during PCI.(11,15) Higher BMI raises attenuation and scatter, prompting automatic exposure-control systems to increase tube output. Obese patients undergoing complex PCI should therefore be flagged before the procedure so that dose-reduction measures are activated from the start.

The score is not intended to replace real-time dose monitoring. Its purpose is to identify, before the procedure, the minority of cases where proactive dose-reduction carries the greatest benefit: the low-risk majority (0.41% RO) needs no change to standard workflow, whereas the 6% high-risk group, with an approximately tenfold higher rate, warrants deliberate action from the outset.

Universal maximal radiation vigilance is the theoretical ideal, but sustained attention across all procedures is not achievable in practice, as routine experience and the literature on operator-level variability both show. Rather than triaging which patients deserve protection, the score addresses where reinforced, attention-intensive measures should be concentrated, complementing rather than replacing baseline ALARA practice. By flagging the minority of procedures that carry most of the risk before irradiation begins, it converts a diffuse and easily eroded vigilance into a targeted, sustainable one.

A high score should prompt defined radiation-sparing behaviours from the outset (low fluoroscopy pulse rate, strict collimation, fewer cine acquisitions, varied working projections, optimized table height and detector position, and dose verbalization at predefined milestones), which cost little prospectively but are far harder to apply once substantial dose has accumulated.

Because the endpoint was rare, the score was constrained to few predictors to avoid overfitting, yielding discrimination appropriate for a pragmatic bedside tool rather than a higher but statistically unreliable estimate from an overparameterized model.

### Limitations

Several limitations deserve emphasis. The study is retrospective and single-center, so unmeasured confounding and local practice patterns may influence the findings. Although equipment and operator composition remained stable throughout the study period, which mitigates two major sources of between-center variability, individual operator-level differences in fluoroscopy habits, angulation preferences and collimation practices were not formally analyzed and may have contributed to residual unexplained variance in radiation exposure. Cumulative AK and fluoroscopy time were available only in subsets of the cohort, reflecting the progressive implementation of automated dose-recording functionality in the catheterization laboratory equipment, procedures performed before 2014 predated the activation of AK logging, rather than selective ascertainment. The composite endpoint therefore denotes exceedance of at least one available threshold, and the observed 0.93% rate is a lower bound on the true prevalence of exceeding any threshold. In ad hoc cases part of the measured dose has been delivered before the score can be computed, so the score stratifies the total session dose rather than the incremental interventional dose. Patient and lesion characteristics were comparable between procedures with and without available AK, supporting the representativeness of the AK subcohort; however, radiation metrics in this subcohort reflect the more recent half of the study period and may not fully capture the radiation environment of the earlier years. Peak skin dose was unavailable and could not be included despite being the most direct metric for skin injury. The composite endpoint was rare, limiting the number of predictors that can be safely included in multivariable logistic models. The score was internally derived and should be considered exploratory until externally validated. Calibration estimates at the upper end of the scale rest on few procedures (12 with a score of 10, six with 11, and one above 11) and are correspondingly uncertain. Male sex was an independent determinant of air kerma but was deliberately not included in the score, to keep it procedurally actionable and centred on modifiable or anticipable procedural factors; adding it increased discrimination only marginally (AUC 0.772 to 0.783) while diluting the low-risk stratum. Discrimination was nonetheless similar in men (AUC 0.780, 95% bootstrap CI 0.736 to 0.824) and in women (0.742, 0.517 to 0.950), the wide interval in women reflecting only 10 events (Supplementary Table 8). Because the composite components had different availability and therefore different denominators, score performance was additionally confirmed on the DAP-only endpoint ascertained in the whole cohort. Finally, procedures were analysed at the procedure level and the anonymised database did not contain a unique patient identifier, so repeated procedures in the same patient could not be accounted for and patient-level cumulative exposure could not be assessed. In addition, the database captured procedural dose indicators but not systematic dermatologic follow-up, so the analysis cannot estimate the incidence of actual radiodermatitis.

## Conclusions

In this real-world cohort of 10,634 consecutive percutaneous coronary interventions, radiation overexposure defined by professional-society alert thresholds occurred in approximately 1% of procedures, five times more often during complex intervention, and the three alert metrics overlapped only partially, indicating that no single metric captures the full radiation burden. Body habitus and procedural complexity independently determined radiation exposure, and a pragmatic integer score based on seven characteristics available after diagnostic angiography identified at-risk procedures with good discrimination and calibration. Because every component of the score is known before the intervention begins, and because the score is implemented in a freely accessible online calculator (https://radar-pci.netlify.app), radioprotective measures and structured post-procedural skin surveillance can be planned in advance for the minority of patients who need them most, rather than reactively once substantial dose has accumulated. External validation in independent cohorts and in contemporary practice is warranted before broader adoption.

## Data Availability

The data that support the findings of this study are not publicly available because they originate from an institutional clinical database and their public deposition was not covered by the ethics approval, but they are available from the corresponding author upon reasonable request. The complete specifications of all models implemented in the online calculator, including all coefficients, are provided in the Supplementary Material. The calculator itself is freely accessible at https://radar-pci.netlify.app.

## Non-standard Abbreviations and Acronyms

PCI: percutaneous coronary intervention
RO: radiation overexposure
CTO: chronic total occlusion
DAP: dose-area product
AK: cumulative reference-point air kerma
BMI: body mass index
AUC: area under the receiver-operating characteristic curve
SIR: Society of Interventional Radiology
CABG: coronary artery bypass grafting
LASSO: least absolute shrinkage and selection operator

